# A booster dose of an inactivated SARS-CoV-2 vaccine increases neutralizing antibodies and T cells that recognize Delta and Omicron variants of concern

**DOI:** 10.1101/2021.11.16.21266350

**Authors:** Bárbara M Schultz, Felipe Melo-González, Luisa F Duarte, Nicolás MS Gálvez, Gaspar A Pacheco, Jorge A Soto, Roslye V Berríos-Rojas, Liliana A González, Daniela Moreno-Tapia, Daniela Rivera-Pérez, Mariana Ríos, Yaneisi Vázquez, Guillermo Hoppe-Elsholz, Omar P Vallejos, Carolina Iturriaga, Marcela Urzua, María S Navarrete, Álvaro Rojas, Rodrigo Fasce, Jorge Fernández, Judith Mora, Eugenio Ramírez, Aracelly Gaete-Argel, Mónica Acevedo, Fernando Valiente-Echeverría, Ricardo Soto-Rifo, Daniela Weiskopf, Alba Grifoni, Alessandro Sette, Gang Zeng, Weining Meng, CoronaVac03CL Study Group, José V González-Aramundiz, Pablo A González, Katia Abarca, Alexis M Kalergis, Susan M Bueno

**Affiliations:** Millennium Institute on Immunology and Immunotherapy, Santiago, Chile; Departamento de Genética Molecular y Microbiología, Facultad de Ciencias Biológicas, Pontificia Universidad Católica de Chile, Santiago, Chile; Departamento de Enfermedades Infecciosas e Inmunología Pediátrica, División de Pediatría, Escuela de Medicina, Pontificia Universidad Católica de Chile, Santiago, Chile; Departamento de Enfermedades Infecciosas del Adulto, División de Medicina, Escuela de Medicina, Pontificia Universidad Católica de Chile, Santiago, Chile; Departamento de Laboratorio Biomédico, Instituto de Salud Pública de Chile; Laboratorio de Virología Molecular y Celular, Programa de Virología, Instituto de Ciencias Biomédicas, Facultad de Medicina, Universidad de Chile, Santiago, Chile; Center for Infectious Disease and Vaccine Research, La Jolla Institute for Immunology (LJI), La Jolla, CA 92037, USA; Department of Medicine, Division of Infectious Diseases and Global Public Health, University of California, San Diego (UCSD), La Jolla, CA 92037, USA; Sinovac Biotech, Beijing, China; Departamento de Farmacia, Facultad de Química y de Farmacia, Pontificia Universidad Católica de Chile, Santiago, Chile; Departamento de Endocrinología, Facultad de Medicina, Escuela de Medicina, Pontificia Universidad Católica de Chile, Santiago, Chile.

**Keywords:** CoronaVac, Phase 3 clinical trial, SARS-CoV-2, COVID-19, inactivated vaccine, Booster dose.

## Abstract

**Background:** CoronaVac^®^ is an inactivated SARS-CoV-2 vaccine approved by the World Health Organization. Previous studies reported increased levels of neutralizing antibodies and specific T cells two- and four-weeks after two doses of CoronaVac^®^, but the levels of neutralizing antibodies are reduced at six to eight months after two doses. Here we report the effect of a booster dose of CoronaVac^®^ on the anti-SARS-CoV-2 immune response generated against variants of concern (VOC) Delta and Omicron in adults participating in a phase 3 clinical trial in Chile.

**Methods:** Volunteers immunized with two doses of CoronaVac^®^ in a four-week interval received a booster dose of the same vaccine between twenty-four and thirty weeks after the 2nd dose. Four weeks after the booster dose, neutralizing antibodies and T cell responses were measured. Neutralization capacities and T cell activation against VOC Delta and Omicron were detected at four weeks after the booster dose.

**Findings:** We observed a significant increase in neutralizing antibodies at four weeks after the booster dose. We also observed an increase in CD4^+^ T cells numbers over time, reaching a peak at four weeks after the booster dose. Furthermore, neutralizing antibodies and SARS-CoV-2 specific T cells induced by the booster showed activity against VOC Delta and Omicron.

**Interpretation:** Our results show that a booster dose of CoronaVac^®^ increases the anti-SARS-CoV-2 humoral and cellular immune responses in adults. Immunity induced by a booster dose of CoronaVac^®^ is active against VOC, suggesting an effective protection.

## Background

The ongoing pandemic caused by severe acute respiratory syndrome coronavirus-2 (SARS-CoV-2) has promoted the rapid development of safe, immunogenic, and effective vaccines against SARS-CoV-2 to be used by the general population, which have successfully reduced the transmission of the disease burden. CoronaVac^®^ is an inactivated SARS-CoV-2 vaccine developed by Sinovac Life Sciences Co., Ltd. (Beijing, China) and is among the current vaccines approved by the World Health Organization (WHO) to combat coronavirus disease 2019 (COVID-19) and one of the most used vaccines worldwide^1, 2^. Phase I/II clinical trials in China demonstrated that this vaccine induces cellular and humoral response upon immunization ^3–5^. Furthermore, the ongoing phase 3 clinical trial in Chile has described an increase in the levels of IgG and neutralizing antibodies in adults aged 18-59 years and ≥ 60 years two- and four-weeks after the second dose of CoronaVac^® 56^. In addition, this vaccination promotes the activation of a T cell immune response against SARS-CoV-2 antigens in a 0-14 immunization schedule ^5^ (two-weeks interval), being an effective vaccine to prevent COVID-19 ^7, 8^. In Chile, 93.7% of the target population has received a first vaccine dose, and 91.4% were fully vaccinated with CoronaVac® on December 10^th^ of 2021 in a 0-28 days vaccination schedule ^9^. Although this primary immunization schedule induces neutralizing antibody present in the serum of vaccinated people ^10^, these titers decrease in time ^6, 11, 12^ and have lower levels of neutralization against highly transmissible variants of concern (VOC) as compared to the original vaccine strain, potentially decreasing the effectiveness of these vaccines as new variants emerge^13–17^. For these reasons, the use of booster doses was approved in Chile in August 2021 in Chile for high-risk populations and adults at five months after administration of the second dose ^18^. In this sense, a report published in October 2021 in Chile, showed that the effectiveness of CoronaVac^®^ against COVID-19 increase from 56% to 80% fourteen days after the application of the booster dose^19^. Notably, a previous study performed in adults aged 18-59 years old demonstrated that a booster dose of CoronaVac^®^, applied six months after the first dose to individuals that previously received two doses of this vaccine, increased the levels of antibodies 3-5-fold as compared to the levels observed four weeks after the second dose ^12^. Here, we further extend these findings by reporting the levels of neutralizing antibodies and specific T cells against SARS-CoV-2 and its activity against VOC Delta and Omicron in adults ≥18 years old that participated in the phase 3 clinical trial carried out in Chile, who were vaccinated in a 0-28-days vaccination schedule and received a booster dose five months after the second dose.

## Materials and methods

### Volunteers and sample collection

Blood samples were obtained from volunteers recruited in the clinical trial CoronaVac03CL (clinicaltrials.gov #NCT04651790) carried out in Chile starting in November 2020. The Institutional Scientific Ethical Committee of Health Sciences reviewed and approved the study protocol at the Pontificia Universidad Católica de Chile (#200708006). Trial execution was approved by the Chilean Public Health Institute (#24204/20) and was conducted according to the current Tripartite Guidelines for Good Clinical Practices, the Declaration of Helsinki ^20^, and local regulations. Informed consent was obtained from all volunteers upon enrollment. Volunteers receive two doses of CoronaVac^®^ (3 µg or 600SU of inactivated SARS-CoV-2 inactivated along with alum adjuvant) in a four-week interval (0-28-day immunization schedule) and then a booster dose five months after the second dose. A complete inclusion and exclusion criteria list have been reported ^5^. On November 11^th^, 2021, one hundred and eighty-six volunteers in the immunogenicity branch received the booster dose. The antibody and cell mediated immune responses were evaluated volunteers that had completed all their previous visits in one of the centers of the study (**Figure 1A**). Blood samples were obtained from all the volunteers before administration of the first dose (pre-immune), two, four, and twenty weeks (or five months) after the second dose, and four weeks after the booster dose (**Figure 1B**).

**Figure 1:**
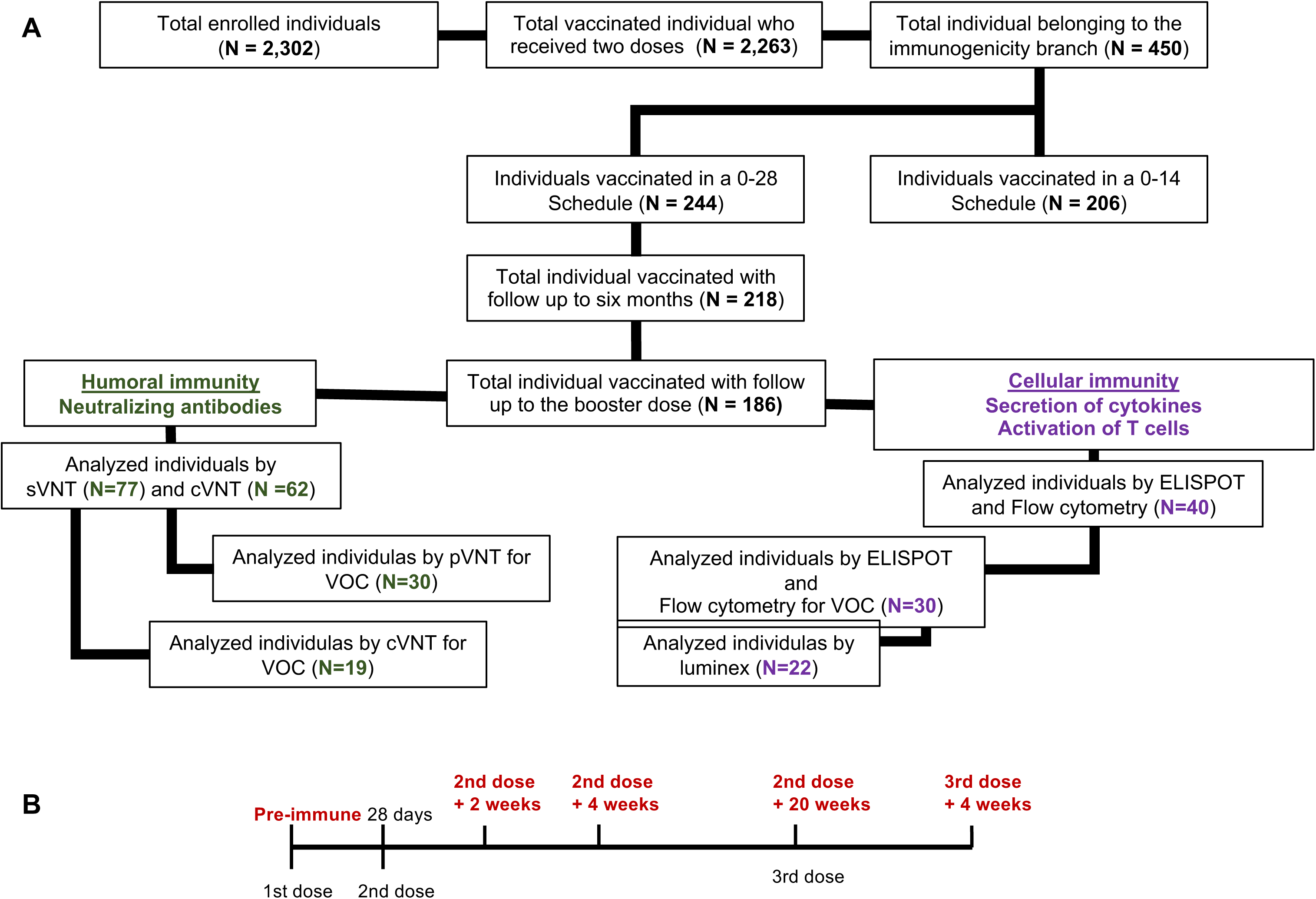
Study profile, enrolled volunteers, and cohort included in the study by November 11^th^, 2021. **A.** From the one hundred and eighty-six vaccinated individuals that received the booster dose, seventy-seven volunteers that received two doses of CoronaVac® in a 28 days interval (0-28 days schedule of vaccination), were selected from the center assigned for the immunogenicity study. Seventy-seven volunteers were tested for neutralizing antibodies by sVNT, sixty-two were selected to analyze neutralizing antibodies by cVNT and forty were selected to analyze cellular immunity. Analyses for immunity against SARS-CoV-2 variants were performed in 30 volunteers for sVNT, pVNT and T cells assays. **B.** Timeline of 0–28 days schedule of vaccination and booster dose immunization. Text in red denotes timepoints at which blood draws occurred.

### Procedures

The presence of antibodies against RBD with neutralizing capacities were measured in sera from seventy-seven volunteers that had completed all their study visits, including one month after the booster dose of CoronaVac^®^. The neutralizing capacities of circulating antibodies were evaluated by a surrogate virus neutralization test (sVNT) (Genscript Cat#L00847-A). Samples were two-fold serially diluted starting at a 4-fold dilution until reaching a 512-fold dilution. Assays were performed according to the instructions of the manufacturer and as reported previously ^5^. Neutralizing antibody titers were determined as the last fold dilution in which the interaction between hACE2 and RBD was inhibited by 30% or more. Samples with a percentage of inhibition ≤30% at the lowest dilution (1:4) were assigned as seronegative with a titer of 2. A sample was considered seropositive when its titer was higher than the pre-immune titer. The percentage of inhibition was determined as: 100 * [OD450nm value of negative control - OD450nm value of sample] / [OD450nm of negative control]. A standard curve was used to plot the neutralization response in the samples as international units (IU) using the WHO International Standard for SARS-CoV-2 antibody (NIBSC code 20/136), which was prepared according to the instructions of the manufacturer ^21^. Data were analyzed using a sigmoidal curve model with a logarithm transformation of the concentration, and the final concentration for each sample was the average of the product of the interpolated IU from the standard curve and the sample dilution factor required to reach the OD450 value that falls within the linear range determined for each sample. Samples with undetermined concentration at the lowest dilution tested (1:4) were assigned to the lower limit of quantification (16.4 IU). The Geometric Mean Units (GMU) and titers (GMT) were represented in **Figure 2** and **Supplementary Figure 1**, respectively. **Table 2** shows comparisons among the visits.

**Table 1:**
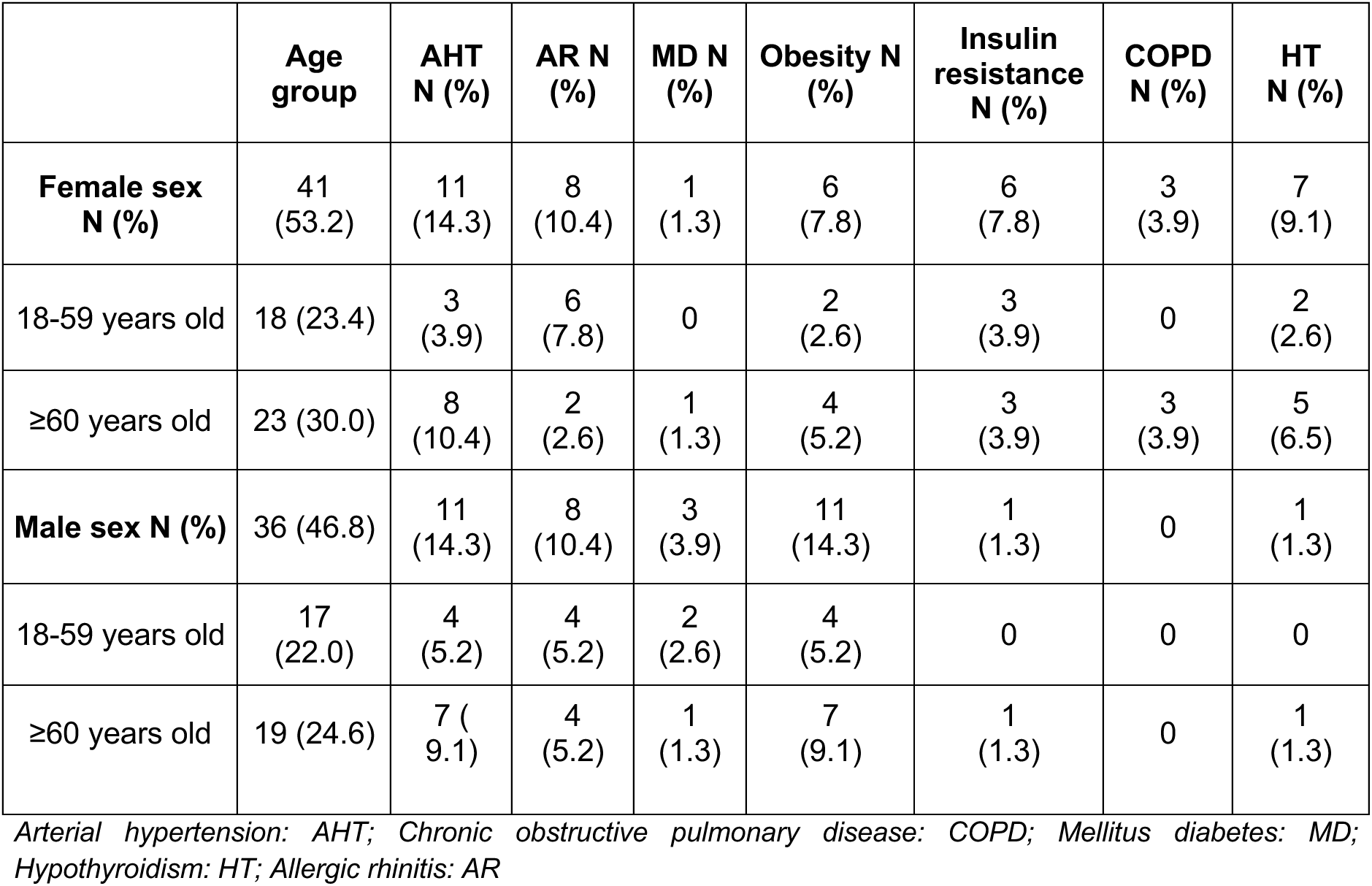
Demographic and clinical data of seventy-seven volunteers analyzed.

**Figure 2:**
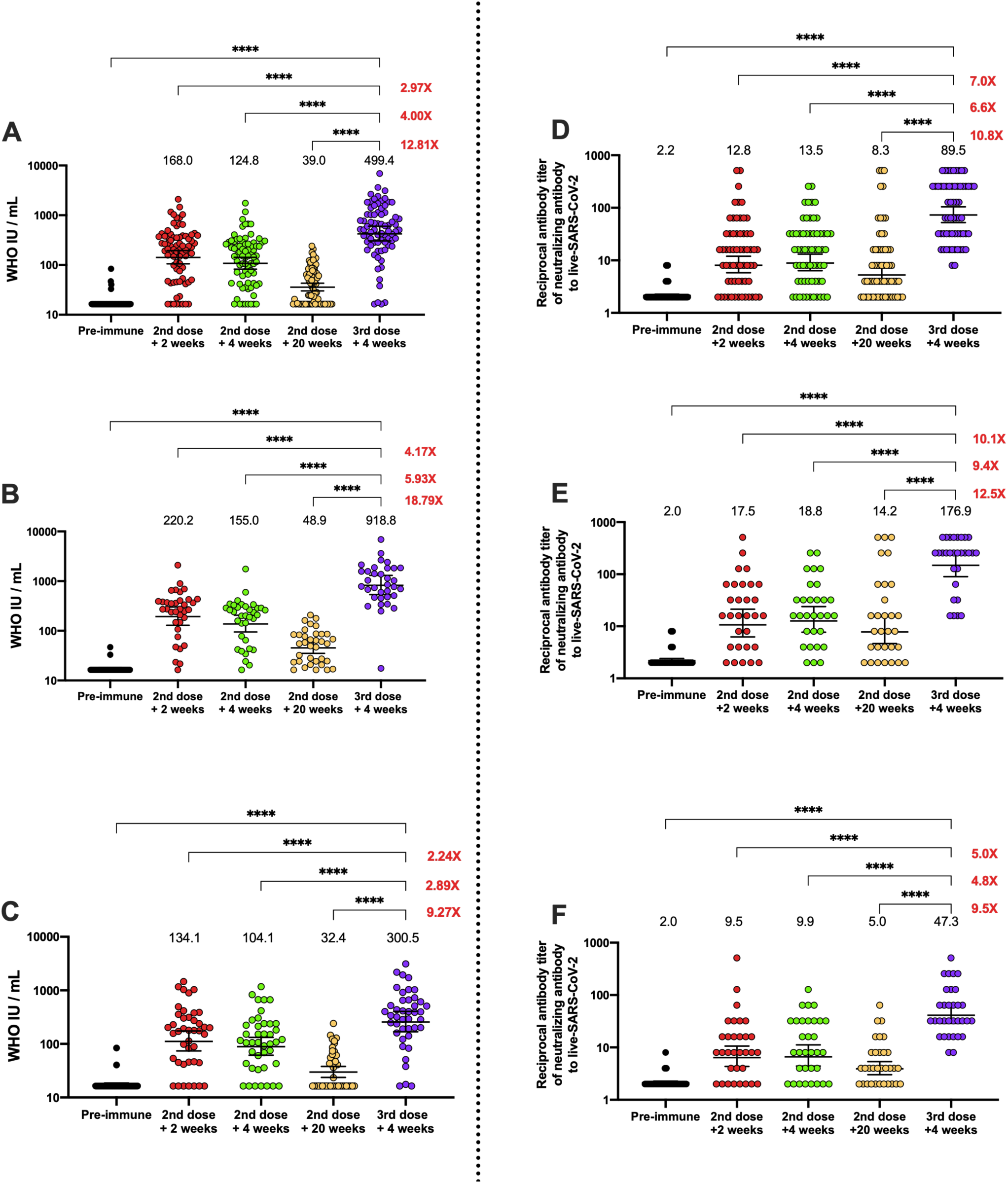
Quantification of circulating antibodies inhibiting the interaction between the S1-RBD and hACE2 and in live SARS-CoV-2 in volunteers that received the booster dose of CoronaVac®. **A-C.** Inhibiting antibodies were detected in serum of volunteers immunized with CoronaVac® using a surrogate Viral Neutralization Test (sVNT), which quantifies the interaction between S1-RBD and hACE2 on ELISA plates. Results were obtained from a total of seventy-seven volunteers (**A**), thirty-six of them were adults between 18-59 years old (**B**), and forty-one of them were ≥ 60 years old (**C**). Data is represented as WHO arbitrary units/mL, the numbers above each set of individual data points show the Geometric Mean Units (GMU), the error bars indicate the 95% CI, and the number at the right represents the fold increase of the GMU four weeks after the third dose as compared with the respective times after administration of the second dose. **D-F**. Neutralizing antibodies were detected in serum of volunteers that received a booster dose of CoronaVac® twenty weeks after the second dose, using a conventional Viral Neutralization Test (cVNT), which quantifies the reduction of cytopathic effect (CPE) in Vero E6 cells infected with SARS-CoV-2. Results were obtained from 62 volunteers (**D**), 30 of them were adults between 18-59 years old (**E**), and 32 of them were ≥ 60 years old (**F**). Data are expressed as the reciprocal of the highest serum dilution preventing 100% cytopathic effect, the numbers above each set of individual data points show the Geometric Mean Titer (GMT), the error bars indicate the 95% CI, and the number at the right represents the fold increase of the GMU the third dose + 4 weeks as compared with the respective times after administration of the second dose. CI were not adjusted for multiplicity and should not be used for inference. A repeated measures One-Way ANOVA test assessed statistical differences to compare all times against 3^rd^ dose + four weeks. ****p<0.0001.

**Table 2:** Seropositivity rates, Geometric Mean Titer (GMT), and Geometric Mean Units (GMU) of circulating neutralizing antibodies against SARS-CoV-2 RBD.

| Methodology | Age group | Indicators | 2<sup>nd</sup> dose + 2 weeks | 2<sup>nd</sup> dose + 4 weeks | 2<sup>nd</sup> dose + 20 weeks | 3<sup>rd</sup> dose + 4 weeks |
| --- | --- | --- | --- | --- | --- | --- |
| **sVNT** | Total Vaccine | Seropositivity n/N | 72/77 | 73/77 | 38/77 | 75/77 |
|  |  | (%) | 93.5 | 94.8 | 49.4 | 97.4 |
|  |  | GMU | 168.0 | 124.8 | 39.0 | 499.4 |
|  |  | 95% CI | 126.8-222.5 | 96.3-161.7 | 32.4-47.0 | 370.6-673.0 |
|  |  | GMT | 25.8 | 16.6 | 3.5 | 53.0 |
|  |  | 95% CI | 19.5-34.2 | 13.1-21.0 | 3.0-4.1 | 40.8-68.8 |
|  | 18-59 | Seropositivity n/N | 35/36 | 36/36 | 24/36 | 36/36 |
|  |  | (%) | 97.2 | 97.2 | 66.7 | 100 |
|  |  | GMU | 220.2 | 155.0 | 48.9 | 918.8 |
|  |  | 95% CI | 150.7-321.7 | 108.0-222.6 | 37.6-63.5 | 623.4-1354 |
|  |  | GMT | 33.3 | 19.1 | 4.3 | 82.8 |
|  |  | 95% CI | 23.4-47.3 | 14.0-26.1 | 3.4-5.4 | 59.7-114.8 |
|  | ≥60 | Seropositivity n/N | 38/41 | 39/42 | 15/42 | 40/42 |
|  |  | (%) | 90.5 | 92.9 | 35.7 | 95.2 |
|  |  | GMU | 134.1 | 104.1 | 32.4 | 300.5 |
|  |  | 95% CI | 89.2-201.6 | 71.8-151.0 | 25.1-41.8 | 203.5-443.6 |
|  |  | GMT | 20.8 | 14.7 | 2.4 | 36.5 |
|  |  | 95% CI | 13.6-31.9 | 10.3-21.0 | 2.4-3.5 | 25.3-52.7 |
| **cVNT** | Total Vaccine | Seropositivity n/N | 49/62 | 51/62 | 44/62 | 62/72 |
|  |  | (%) | 79.0 | 82.3 | 71.0 | 100 |
|  |  | GMT | 12.8 | 13.5 | 8.3 | 89.5 |
|  |  | 95% CI | 8.8-18.5 | 9.6-19.2 | 5.6-12.2 | 64.0-125.2 |
|  | 18-59 | Seropositivity n/N | 25/30 | 27/30 | 23/30 | 30/30 |
|  |  | (%) | 83.3 | 90.0 | 76.7 | 100 |
|  |  | GMT | 17.5 | 18.8 | 14.2 | 176.9 |
|  |  | 95% CI | 9.8-31.3 | 11.2-31.7 | 7.1-28.4 | 111.7-280.1 |
|  | ≥60 | Seropositivity<br>n/N | 24/32 | 24/32 | 21/32 | 32/32 |
|  |  | (%) | 75.0 | 75.0 | 65.6 | 100 |
|  |  | GMT | 9.5 | 9.9 | 5.0 | 47.3 |
|  |  | 95% CI | 5.8-15.4 | 6.2-15.8 | 3.5-7.0 | 32.1-69.5 |

sVNT: Surrogate Virus Neutralization; cVNT: Conventional Virus Neutralization; GMT: Geometric mean titer; GMU: Geometric mean units.

Conventional virus neutralization tests (cVNT) were performed in sixty-two of the previous seventy-seven volunteers and evaluated as previously reported ^5^. Briefly, Vero E6 cells were infected with a SARS-CoV-2 strain obtained by viral isolation in tissue cultures (33782CL-SARS-CoV-2 strain, D614G variant). Neutralization assays were carried out by the reduction of cytopathic effect (CPE) in Vero E6 cells (ATCC CRL-1586). The titer of neutralizing antibodies was defined as the highest serum dilution that neutralized virus infection, at which the CPE was absent as compared with the virus control wells (cells with CPE). Vero E6 cells were seeded in 96-well plates (4×10^4^ cells/well). For neutralization assays, 100 µL of 33782CL-SARS-CoV-2 (at a dose of 100 TCID_50_) were incubated with serial dilutions of heat-inactivated sera samples (dilutions of 1:4, 1:8, 1:16, 1:32, 1:64, 1:128, 1:256, and 1:512) from participants for 1h at 37 °C. Cytopathic effect on Vero E6 cells was analyzed 7 days after infection.

A pseudotyped virus neutralization test (pVNT) assay was performed to assess the capacity of the antibodies against SARS-CoV-2 VOC in samples from thirty volunteers of the seventy-seven previously analyzed by sVNT. As previously reported ^14^, a HIV-1 backbone expressing firefly luciferase as a reporter gene and pseudotyped with the SARS-CoV-2 spike glycoproteins (HIV-1-SΔ19) from from lineage B.1 (D614G) or variants Delta (T19R, del157/158, L452R, T478K, D614G, P681R, D950N) and Omicron (A67V, ΔH69-V70, T95I, Y145D, ΔG142 -V143-Y144, ΔN211, EPE 213-214, G339D, S371L, S373P, S375F, K417N, N440K, G446S, S477N, T478K, E484A, Q493R, G496S, Q498R, N501Y, T547K, D614G, H655Y, N679K, P681H, N764K, N865K, Q954H, N969K, L981F) was prepared as previously described ^22^. Serum samples were two-fold diluted starting at 1:10 or 1:4 and the estimation of the ID80 was obtained using a 4-parameter nonlinear regression curve fit measured as the percent of neutralization determined by the difference in average relative light units (RLU) between test samples and pseudotyped virus controls. Also, cVNT assays were performed to assess the capacity of the antibodies against SARS-CoV-2 Delta variant in samples from nineteen volunteers of the seventy-seven previously analyzed by sVNT.

ELISPOT and flow cytometry assays were performed to evaluate the cellular immune response in forty volunteers of the seventy-seven previously analyzed (**Figure 1A**), stimulating PBMCs with four Mega Pools (MPs) of peptides derived from the proteome of SARS-CoV-2 ^23^: peptides from the S protein of SARS-CoV-2 (MP-S), the remaining proteins of the viral particle (excluding S protein peptides) (MP-R), and shorter peptides from the whole proteome of SARS-CoV-2 (MP-CD8-A and MP-CD8-B) ^23^. Thirty of the previously analyzed volunteers were also stimulated with three Mega Pools (MP) of VOC, provided by La Jolla Institute for Immunology. MP derived from the S protein of SARS-CoV-2 WT, SARS-CoV-2 B1.617.2 MP 4326 (Delta variant), and SARS-CoV-2 B1.1.529 MP 4359 (Omicron variant) ^14^ were used to evaluate T cell activation at four weeks after the booster dose. Positive and negative controls were held for each assay. The number of Spot Forming Cells (SFC) for interferon gamma (IFN-γ) were determined by ELISPOT and the expression of Activation-Induced Markers (AIM) by T cells was evaluated by flow cytometry. Assays were performed according to the instructions of the manufacturer and as reported previously ^5^. Further details on the ELISPOT assay, antibodies used for flow cytometry, and the respective protocols can be found in the Supplementary information and **Supplementary Table 1**.

Interleukin 2 (IL-2) and IFN-γ secretion were evaluated in the supernatants of twenty-two volunteers previously stimulated for 20h with SARS-CoV-2 MP of peptides derived from the Spike protein of VOC, using a Luminex 200 xMap multiplex system (Luminex Corporation, Austin, TX). The limit of detection for the cytokine measured ranged from 4.2 to 13,390 pg/mL, according to manufacturer’s instructions. Further experimental details can be found in **Supplementary Information**.

### Statistical analyses

Statistical differences for the immunogenicity results considered repeated measures ANOVA with the Geisser-Greenhouse correction and Dunnet’s *a posteriori* multiple tests to compare between the booster dose and the other visits. Analyses were performed over the base 10 logarithms of the data for neutralizing antibody by sVNT, cVNT and pVNT. Cellular immune responses were analyzed by a Friedman test for repeated measures for ELISPOT and flow cytometry for the comparisons between booster dose and the other visits. Secretion of cytokines were compared between the secretion induced by the WT strain against the VOC Delta and Omicron by repeated measures ANOVA. The significance level was set at 0.05 for all the analyses. All data were analyzed with GraphPad Prism 9.0.1.

## Results

### A booster dose of CoronaVac^®^ induces a significant increase in antibody titers with neutralizing capacity in adults

One hundred and eighty-six volunteers from the immunogenicity branch that received a booster dose of the CoronaVac^®^ were included in this study. The first dose of the vaccine was inoculated from January to March of 2021, and the second dose was inoculated 28 days after the first one. The neutralizing capacity of serum antibodies was evaluated in seventy-seven and sixty-two volunteers by sVNT and cVNT, respectively, at the five different time-points indicated in **Figure 1B**.

As shown in **Figures 2A and D**, the peak level of antibodies with neutralizing capacity in the total population evaluated, tested by sVNT and cVNT, is reached at two weeks after the second dose (GMU 168.0, 95% CI=19.5-34.2 and GMT, 95% CI=) and four weeks after the second dose (GMU 124.8, 95% CI=96.3-161.7 and GMT 13.5, 95% CI=9.6-19.2). However, this neutralizing capacity significantly decreased twenty weeks after the second dose (GMU 39.0, 95% CI=32.4-47.0 and GMT 8.3, 95% CI=9.6-19.2), which is in line with previous reports where the immunity against SARS-CoV-2 wanes six months after infection or vaccination ^24, 25^. After the booster dose, the neutralizing capacity of the antibodies increased even more than the one reported two weeks after the second dose (GMU 499.0, 95% CI=370.6-673.0 and GMT 89.5 ± 64.0-125.2). Overall, we observed that four weeks after the booster dose the neutralizing capacity increased more than 12-fold (sVNT) and 10-fold (cVNT) as compared to the response at twenty weeks after the second dose, and almost 3-fold as compared to two weeks after the second dose (**Figures 2A and D**).

In adults 18-59 years old, the neutralizing capacity of circulating antibodies tested by sVNT and cVNT (**Figures 2B and E**, respectively) reached its maximum four weeks after the booster dose (GMU 918.8, 95% CI=623.4-1354 and 176.9, 95% CI=111.7-280.1) increasing more than 18- and 12-fold as compared to five months after the second dose (GMU 48.9, 95% CI=37.6-63.5 and GMT 14.2, 95% CI=7.1-28.4) and more than 4-fold as compared to two weeks after the second dose (GMU 220.2, 95% CI=150.7-321.7 and GMT 17.5, 95% CI=9.8-31.3) (**Figures 2B and E**). The seropositivity rate in this group reached 100% at four weeks after the booster dose (**Table 2**). On the other hand, 53.2% of the total volunteers analyzed here were adults ≥60 years. In this group, the same tendency was observed, as seen in **Figure 2C and F**, observing an increase in the level of neutralizing antibodies evaluated by both techniques of more than 9-fold at four weeks after the booster dose (GMU 300.5, 95% CI=203.5-443.6 and GMT 47.3, 95% CI=32.1-69.5) as compared to the response observed twenty weeks after the second dose (GMU 32.4, 95% CI=25.1-41.8 and GMT 5.0, 95% CI=3.5-7.0). Equivalent to the 18-59 years old group, the seropositivity rate in this age group reached 100% four weeks after the booster dose (**Table 2**). The seropositivity rate achieved at four weeks after the booster dose was the highest when compared with the other visits in this study in the total vaccinated group and in both groups analyzed.

### A booster dose of CoronaVac^®^ induces a robust cellular immune response in adults

The cellular responses following a booster dose of CoronaVac^®^ were evaluated in 40 volunteers. We observed that CD4^+^ T cell activation was increased twenty weeks after the second dose as compared to the other time points in both age groups, suggesting that CoronaVac® can stimulate CD4^+^ T cell responses that are sustained over time (**Figure 3A-C**). Importantly, we observed a significantly further increase in CD4^+^ T cell activation in both age groups following the booster dose, as compared to the pre-immune sample and samples obtained at two and four weeks after the second dose (**Figure 3A-C**). However, this difference was not significant as compared to the sample obtained twenty weeks after the second dose (**Figure 3A-C**). Moreover, we did not observe a significant increase in the expression of AIM by CD8^+^ T cells following the booster dose as compared to any other time point, suggesting that specific CD8^+^ T cell responses induced by CoronaVac^®^ are not detected with the current methodologies, even after a third dose (**Supp. Figure 2A and C**). Accordingly, we observed an increase in IFN-γ production upon stimulation with mega-pools of peptides (MPs) S and R by ELISPOT at four weeks after the booster dose for both groups, as compared to the pre-immune sample (**Figure 3D-F**). As in the case of flow cytometry, we did not observe significant increase of IFN-γ^+^ SFCs upon stimulation with CD8 MPs at any time point (**Supp. Figure 2**). These results suggest that although humoral responses decrease over time following vaccination with CoronaVac^®^, CD4^+^ T cell responses remain significantly increased as compared to pre-immune samples and the booster dose promotes a small increase both IFN-γ production and CD4^+^ T cell activation that is not significantly different as compared to the levels observed 20 weeks after the second dose.

**Figure 3:**
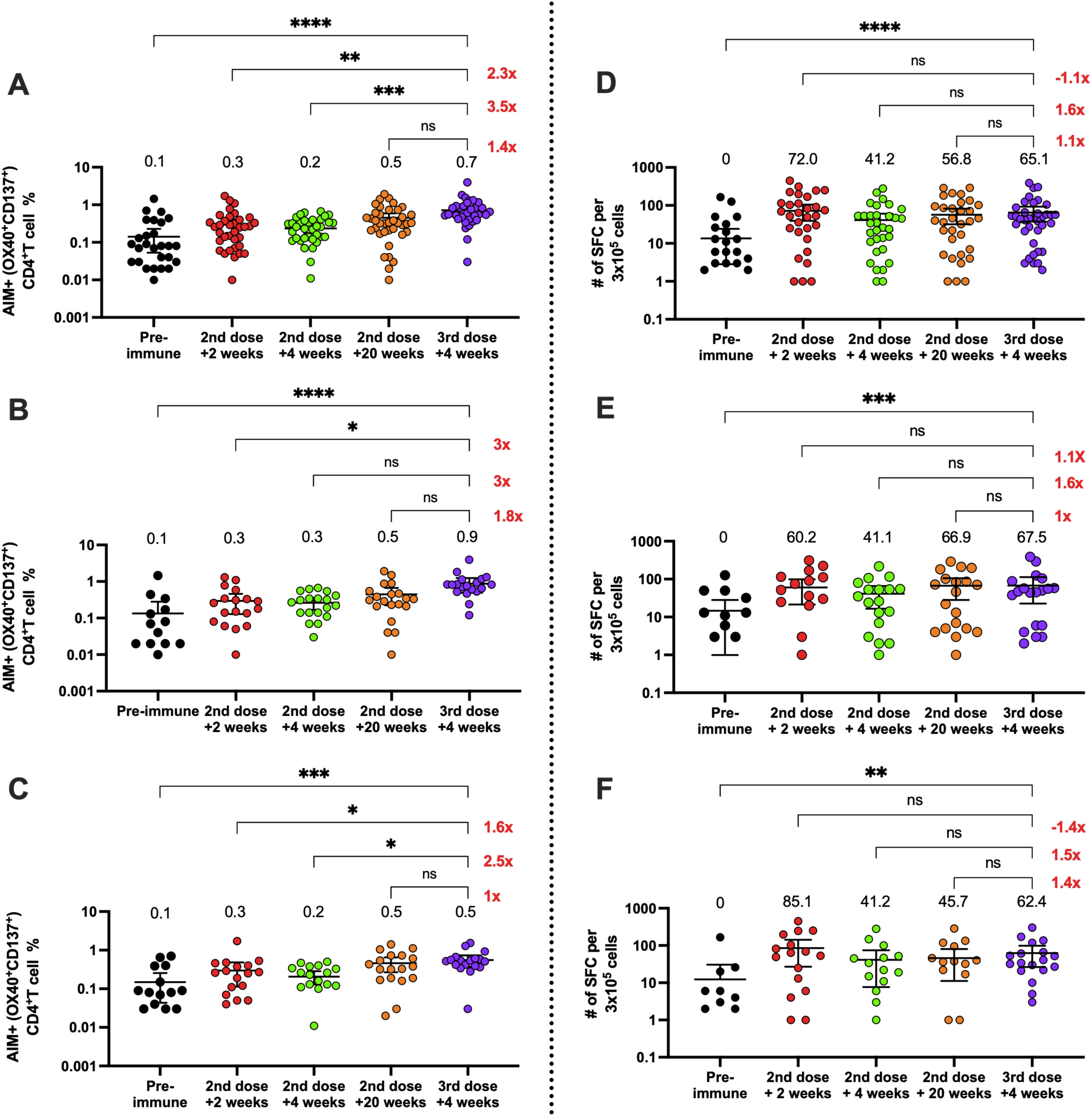
Changes in activation-induced markers (AIMs) expression in CD4^+^ T cells and in the number of IFN-γ-secreting cells specific for SARS-CoV-2 after a booster dose of CoronaVac^®^. **A-C.** AIM^+^ CD4^+^ T cells were quantified in peripheral blood mononuclear cells of volunteers that received a booster dose of CoronaVac® twenty weeks after the second dose by flow cytometry, upon stimulation with mega-pools of peptides derived from SARS-CoV-2 proteins. The percentage of activated AIM^+^ CD4^+^ T cells (OX40^+^, CD137^+^) were determined upon stimulation for 24h with MP-S+R in samples obtained at pre-immune, two weeks after the second dose, four weeks after the second dose, twenty weeks the second dose, and four weeks after the booster dose. Data from flow cytometry was normalized against DMSO and analyzed separately by a Friedman test against the booster dose. Results were obtained from a total of forty volunteers (**A**), twenty-one were of them were adults between 18-59 years old (**B**), and nineteen of them were ≥ 60 years old (**C**). Changes in the secretion of IFN-γ were quantified as the number of Spot Forming Cells (SFCs) in peripheral blood mononuclear cells of volunteers that received a booster dose of CoronaVac® 20 weeks after the second dose. **D-F.** Data was obtained upon stimulation with MP-S+R for 48h in samples obtained at pre-immune, two weeks after the second dose, four weeks after the second dose, twenty weeks the second dose, and four weeks after the booster dose. Results were obtained from a total of 40 volunteers (**D**), 21 were of them were adults between 18-59 years old (**E**), and 19 of them were ≥ 60 years old (**F**). Data from ELISPOT were analyzed separately by Friedman test against the booster dose *p<0.05; **p<0.005; ***p< < 0.001; ****p<0.0001.

### Neutralizing antibodies and specific T cells induced by a booster dose of CoronaVac® recognize Delta and Omicron variants of SARS-CoV-2

As we observed that the neutralization capacity and the T cell responses increased significantly with the booster dose and knowing that vaccinated volunteers exhibit decreased neutralization against VOC^14^, we proceeded to evaluate the neutralizing capacities of antibodies in the serum from thirty booster-vaccinated individuals in pseudotyped virus neutralization test (pVNT) assay against two variants of concern of SARS-CoV-2, comparing with the level obtained for the D614G SARS-CoV-2 variant (B.1 lineage, **Figure 4A-B**). We observed that the titers of antibodies with neutralizing capacities against Delta and Omicron variant show a significant reduction as compared to the levels achieved for the D61G variant (D614G: GMT 241.8, CI=155.7-375.6, Delta: 159.2, CI=99.1-256.0 and Omicron: GMT 50.7, CI=30.4-84.8), with a reduction of 1.5 for Delta and 4.8 for Omicron (**Figure 4A**). However, when we compared the changes in seropositivity for Delta and Omicron (**Figure 4B**), we observed a 93% and 76.7%, respectively, following the booster dose (**Table 3**). Neutralization assays against Delta variant with a cVNT in a different group of nineteen volunteers also show that antibodies induced four weeks after the booster dose have reduced capacity to neutralize this VOC (**Supp Figure 4A**), although the seropositivity rate observed is 84% (**Supp Figure 4B**).

**Figure 4:**
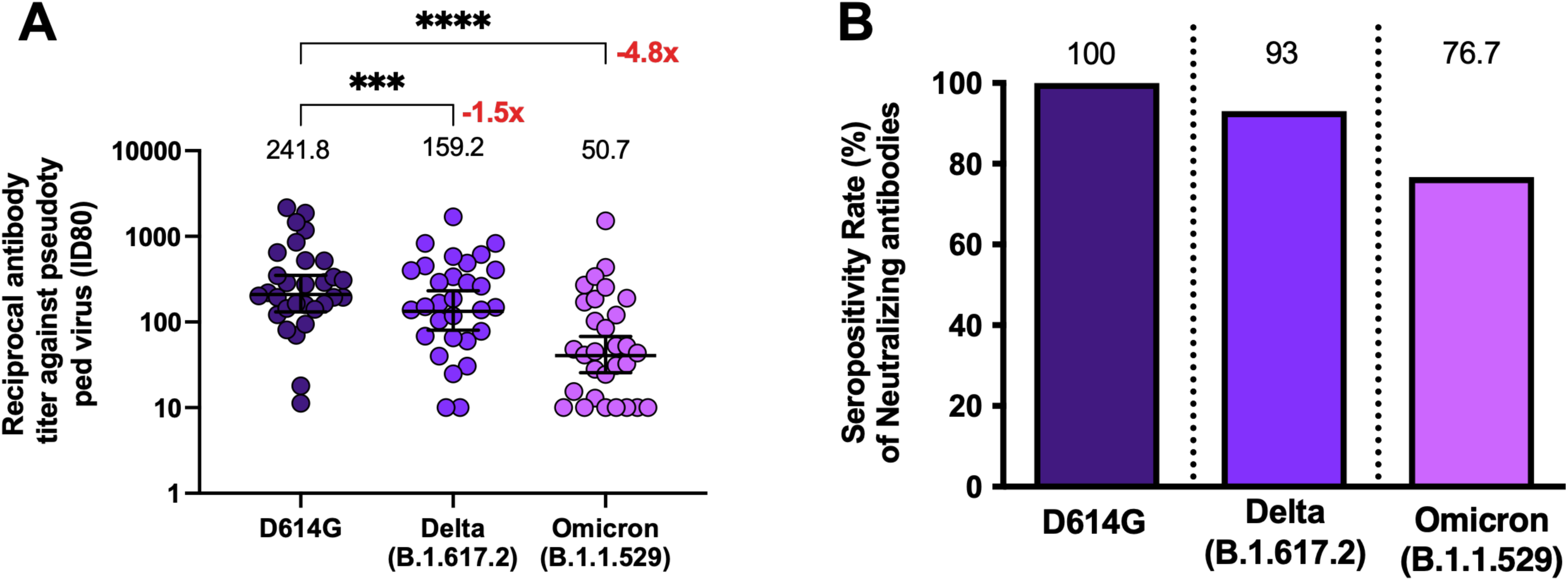
Quantification of circulating neutralizing antibodies against SARS-CoV-2 variants in volunteers that received the booster dose of CoronaVac®. **A.** Neutralizing antibodies were detected in the serum of thirty volunteers, four weeks after the booster dose of CoronaVac®, using a pseudotyped virus neutralization test (pVNT). Data are expressed as the reciprocal of the highest dilution preventing 80% of the infection (ID80). Numbers above the bars show the Mean, and the error bars indicate the 95% CI. The number at the right represents the fold decrease of the GMT four weeks after the booster dose as compared with the response of D614G **B**. Seropositivity rate of neutralizing antibodies is shown for each time point analyzed. Numbers above the bars show the percentage of seropositivity rate in the respective graphs. Numbers above the bars show the percentage of seropositivity rate in the respective graphs. A repeated measures One-Way ANOVA test assessed statistical differences of the GMT to compare each variant against D614G. *p<0.05; ***p < 0.001; ****p<0.0001

**Table 3:** Seropositivity rates, Geometric Mean Titer (GMT) of circulating neutralizing antibodies against SARS-CoV-2 RBD of D614G and variants of concern (Delta and Omicron).

|  | Variant | D614G | Delta<br>(B.1.617.2) | Omicron<br>(B.1.1.529) |
| --- | --- | --- | --- | --- |
| **pVNT** | Indicators | 3rd dose<br>+ 4<br>weeks | 3rd dose + 4<br>weeks | 3rd dose + 4<br>weeks |
|  | Seropositivity<br>n/N | 30/30 | 28/30 | 23/30 |
|  | (%) | 100 | 93.3 | 76.6 |
|  | GMT | 241.8 | 159.2 | 50.7 |
|  | 95% CI | 155.7-375.6 | 99.1-256.0 | 30.4-84.8 |

*GMT: Geometric mean titer.*

The cellular responses for VOC following a booster dose of CoronaVac^®^ were also evaluated in thirty volunteers using MPs of peptides derived from the Spike protein of Delta and Omicron variants. We observed equivalent numbers of AIM by CD4^+^ T cells after four weeks of the booster dose upon stimulation with MP-S of SARS-CoV-2 WT, Delta, or Omicron variant (**Figure 5A**), with no significant differences between the response against the MP-S of the variants as compared to the MP-S of the WT strain. IFN-γ secreting T cells were also analyzed in these samples and no differences were observed **(Figure 5B)**. We also quantified the production of different cytokines in the supernatant of PBMCs stimulated with the MP-s of WT, Delta, and Omicron variants, observing that at four weeks after the booster dose the stimulated cells secrete equivalent levels of IL-2 and IFN-γ (**Figure Supp 3**). These results suggest that although the humoral response measured as neutralization capacities and seroconversion against these VOC is lower as compared to the humoral response against the D614G strain, the cellular responses against SARS-CoV-2 VOC is equivalent to the responses elicited by the wild type strain in volunteers vaccinated with booster dose.

**Figure 5:**
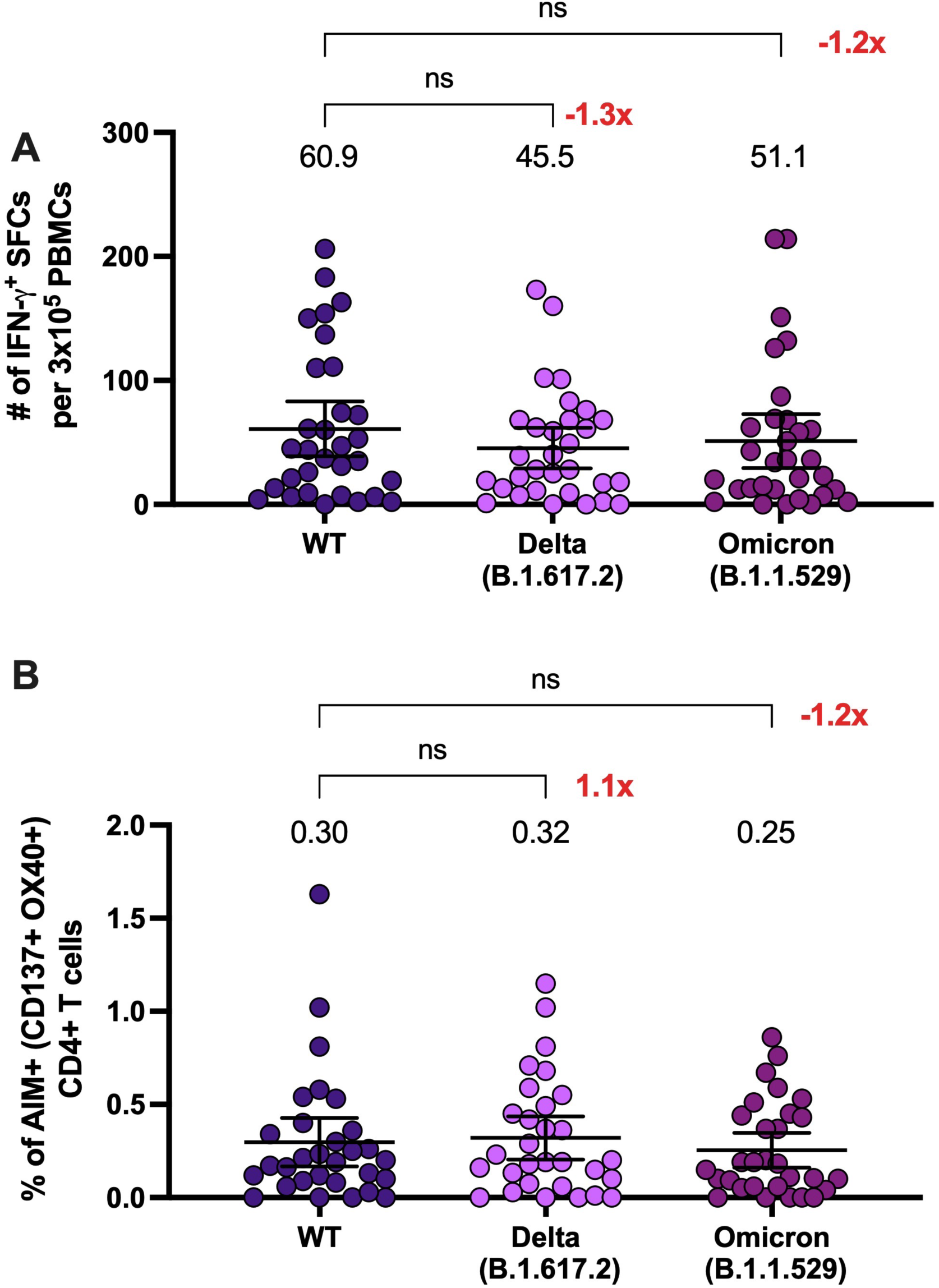
A booster dose of CoronaVac^®^ induce changes in the number of IFN-γ-secreting cells and in activation-induced markers (AIMs) expression in CD4^+^ T cells specific for the Spike protein of SARS-CoV-2 variants. **A.** Changes in the secretion of IFN-γ, determined as the number of Spot Forming Cells (SFCs) were determined. Data was obtained upon stimulation of PBMC with MP-S of variant of concern of SARS-CoV-2 for 48h in samples obtained four weeks after the booster dose. Data shown represent mean + 95%CI. Data from thirty volunteers were analyzed at four weeks after the booster dose to compare among the MP-S of the variant of concern. Data from ELISPOT were analyzed separately by Friedman test against the WT MP-S. No significant differences were obtained. **B.** AIM^+^ CD4^+^ T cells were quantified in peripheral blood mononuclear cells of thirty volunteers four weeks after that received a booster dose of CoronaVac® by flow cytometry, upon stimulation with mega-pools of peptides derived from proteins of variant of concern of SARS-CoV-2. The percentage of activated AIM^+^ CD4^+^ T cells (OX40^+^, CD137^+^) were determined upon stimulation for 24h with MP-S+R in samples obtained four weeks after the booster dose. Data shown represent mean + 95%CI. Data from flow cytometry was normalized against DMSO. No significant differences were obtained between WT and the variant MP stimulation.

## Discussion

In this report we evaluated the humoral and cellular immune response generated four weeks after the application of a booster dose of inactivated CoronaVac® vaccine in a cohort of volunteers enrolled in the phase 3 clinical trial held in Chile. The data reported here showed that although there was an adequate humoral response after two doses of CoronaVac^®^, with a 65.9% of effectiveness in preventing COVID-19 ^8^, both the sVNT and cVNT assays showed a decrease in the GMT of neutralizing capacities of circulating antibodies against SARS-CoV2 twenty weeks after the second dose (**Figure 2**). Due to this decrease in neutralizing capacities, a booster dose of CoronaVac^®^ was evaluated in a clinical study in China, showing promising results in enhanced humoral immune responses^12, 26^. The evaluation of the immune response reported here shows that after the booster dose, the neutralizing titers and seroconversion rates increase in the whole group, to a higher extent than two weeks after the second dose, where the peak in neutralization was previously observed, which is in line with the observed by *Clemens et a*l.,^15^ . Also, we observed a steady activation of the CD4^+^ T cells and secretion of IFN-γ during the time-points evaluated (**Figure 3**).

Since the neutralizing antibody titers correlated with protection against SARS-CoV-2 infection ^10^, these results likely imply a better outcome and protection against COVID-19, as reported in previous studies performed in Israel that showed a decrease in the transmission and the disease severity disease by this virus twelve or more days after booster inoculation. In Chile, the effectiveness and prevention in hospitalization increased when assessed fourteen days after the booster dose of CoronaVac^® 19, 27^. Another study, performed with a booster dose of CoronaVac^®^, showed that an additional dose result in good neutralization capacity against parental SARS-CoV-2 and against Delta variant four weeks after the booster dose, generating a long-lasting humoral response, which was due to an enhancement of the memory immune response generated by B cells ^26^.

Adults ≥60 years old produced lower levels of antibodies with neutralizing capacities than the whole group during this study, which was also described previously (**Figure 2C and F**) ^5^. This result is in line with previous data reported for a population vaccinated in Chile ^6^, among hospital workers who received two doses of CoronaVac^® 28^, and in a study with the mRNA-1273 vaccine ^29^. In this sense, our results are equivalent to those described in a phase I/II of the clinical trial performed with CoronaVac^®^ in China, showing that the neutralizing antibody titers in this group decreased at five months after the second dose and that a booster dose is required 6-8 months after the first vaccination to rapidly increase and maintain the neutralizing antibody titers ^30^.

In the case of the T cell response (**Figure 3**), other studies have shown that Pfizer BNT162b2 and mRNA-1273 induce durable CD4^+^ T cell activation and cytokine production up to six months following vaccination, but it remains to be elucidated whether expression of AIM by CD4^+^ T cells and cytokine production further increase following a booster dose with these vaccines ^31, 32^. Here, we observed that the activation of CD4^+^ T cells and IFN-γ production stays increased up to twenty weeks after the second dose, and after the booster dose both parameters increased in the 18-59 years old group and was maintained at the levels observed twenty weeks after the second dose in adults ≥60 years old. In contrast to BNT162b2 and mRNA-1273 vaccines, CoronaVac^®^ delivers not only the Spike protein upon immunization but also other viral antigens, which may explain why vaccinated individuals still display AIM^+^ CD4^+^ T cells five months after the second dose, regardless of a third dose.

When the neutralization capacity analyzed by pVNT of the VOC Delta and Omicron was evaluated four weeks after the booster dose, we observed differences in the neutralization capacity as compared to the D614G variant (lineage B.1), which does not exhibit mutations in the RBD of the S1 protein (**Figure 4 and table 3**). We previously reported that CoronaVac^®^ is able to induce neutralization against the Delta variant at 4 weeks after the second dose, although to a lesser extent compared to the WT strain ^14^. Another study recently reported a significant increase in the neutralizing capacity after a booster dose with CoronaVac^®^ for the Delta variant, as compared to the levels observed in volunteers vaccinated with two doses ^26, 33^. Although we did not observe similar levels of enhanced neutralization against the variants Delta after the booster dose using pVNT (**Figure 4A**), the seropositivity against Delta variant is almost 100% (**Figure 4B**)^33^. Here, we also show that a booster dose induces neutralization against the variant Omicron, which has rapidly spread worldwide and is the predominant circulating variant to date ^34^. The high number of mutations described in the RBD of this variant has been associated with increased evasion of neutralizing responses in either unvaccinated or vaccinated subjects ^34, 35^. Although the neutralization observed in subjects vaccinated with a booster dose of CoronaVac^®^ is significantly lower to the observed against the D614G variant, we observed a seropositivity of 76.7% following the booster, suggesting some degree of protection in most of the vaccinees. In this sense, it has been reported that a heterologous schedule of vaccination may induce a higher neutralization ability and a better neutralization against variants of concern as Delta ^36^ and Omicron^16^. In line with this, a heterologous vaccination with CoronaVac^®^ and a booster dose of Pfizer BNT162b2 showed a good neutralization titer against VOC Delta and Omicron, with respect to the ancestral virus ^16^. Similarly, a comparison between heterologous and homologous booster schedules after the vaccination with CoronaVac^®^, shows an increase in neutralization against the VOC Delta and Omicron^15^. There are discrepancies between the results in neutralization titers, which can be attributed to the neutralization assays performed and/or the study population; however, important booster responses are observed in these studies, and seropositivity reached after the booster dose of CoronaVac^®^ against VOC are also similar ^16^.

In the case of the cellular response, this is the first report to characterize CD4^+^ T cell responses following a booster dose of CoronaVac^®^ against the Omicron variant of SARS-CoV2. Previous studies using the same MP from VOC evaluated here have shown that CD4^+^ T cells respond to VOC in a similar extent as compared to the ancestral strain in individuals vaccinated with CoronaVac^® 14, 37^ and mRNA vaccines, which has been explained by the high conservation of T cell epitopes. In this sense, the booster dose of CoronaVac^®^ induces the expression of CD4^+^ T cell activation markers and secretion of IFN-γ and IL-2 against the VOC Delta and Omicron, which is comparable to the response generated against the WT strain (**Figure 5**). In line with this, a recent study has shown that T cell responses against the ancestral strain are cross-reactive against the Omicron variant in convalescent individuals and volunteers vaccinated with Pfizer BNT162b2 ^38^, supporting the idea that the induction of T cell responses against the ancestral strain may be protective against the Omicron variant.

Our report shows that the booster dose of CoronaVac^®^ in a 0-28 days schedule induces antibodies with neutralizing capacities, which are higher than the levels observed at 2- and 4-weeks after the second dose, generating an increased humoral response even in adults ≥60 years old. Besides this, our results suggest that a third dose of CoronaVac^®^ supports CD4^+^ T cell activation, which may confer either protection or enhanced immune responses against the virus and prevent severe disease following exposure to SARS-CoV-2 exposure. Importantly, the humoral and cellular immune response promoted by a booster dose of CoronaVac shows activity against Delta and Omicron variants and probably results in better effectiveness of this vaccine during predominance of these VOC.

### Limitations

This study presents several limitations, such as the reduced sample size for the assays and the absence of data for neutralization against Omicron variant with a conventional viral neutralization test. The assessment of total antibody response against Spike proteins and other SARS-CoV-2 proteins would also add additional information about the humoral immune response against SARS-CoV-2 after the booster dose. Due to the limit of quantification of the technique, samples with undetermined concentration at the lowest dilution tested (1:4) were assigned the lower limit of quantification (16.4 IU) and other neutralization assays.

## Data Availability

All data produced in the present study are available upon reasonable request to the authors

## Acknowledgments

We would like to thank the support of the Ministry of Health, Government of Chile; Ministry of Science, Technology, Knowledge, and Innovation, Government of Chile; The Ministry of Foreign Affairs, Government of Chile, and the Chilean Public Health Institute (ISP). We also would like to thank Rami Scharf, Jessica White, Jorge Flores and Miren Iturriza-Gomara from PATH for their support on experimental design and discussion. Alex Cabrera and Sergio Bustos from the Flow Cytometry Facility at Facultad de Ciencias Biológicas, Pontificia Universidad Católica de Chile for support with flow cytometry. We also thank the Vice Presidency of Research (VRI), the Direction of Technology Transfer and Development (DTD), the Legal Affairs Department (DAJ) of the Pontificia Universidad Católica de Chile. We are also grateful to the Administrative Directions of the School of Biological Sciences and the School of Medicine of the Pontificia Universidad Católica de Chile for their administrative support. We would also like to thank the members of the independent data safety monitoring committee (members in the SA) for their oversight, and the subjects enrolled in the study for their participation and commitment to this trial. This project has been funded in whole or in part with Federal funds from the National Institute of Allergy and Infectious Diseases, National Institutes of Health, Department of Health and Human Services, under Contract No. Contract No. 75N9301900065 to A.S, A.G. and D.W.

Note: Members of the CoronaVac03CL Study Group are listed in the Supplementary Appendix (SA).

