## Supplementary Information for "A booster dose of an inactivated SARS-CoV-2 vaccine increases neutralizing antibodies and T cells that recognize Delta and Omicron variants of concern"

### Materials and methods

#### Study design, volunteers, and vaccine

The clinical trial (clinicaltrials.gov NCT04651790) was conducted in Chile at eight different sites, six located in Santiago city (Metropolitan Region) and two in the V Region of Valparaiso. The committee members that reviewed and approved this trial protocol (Institutional Scientific Ethical Committee of Health Sciences, Pontificia Universidad Católica de Chile, Approval #200708006) were: Claudia Uribe PUC/Committee president; Colomba Cofré/Committee Vice-president; Andréa Villagrán/Executive secretary; Jorge Muñoz/External Lawyer; Gustavo Kaltwasser/External member; Alysa Garay/Community representative; Marisa Torres/Public Health Department; Carolina Méndez/Speech therapy representative; Luis Villarroel/Public Health Department; Pablo Brockman/Respiratory Diseases in Children Department. Website: http://eticayseguridad.uc.cl/comite-etico-cientifico-facultad-de-medicina-uc.html).

#### Procedures

For the isolation of sera, 20 mL of blood were collected in anticoagulant tubes and distributed in 2 tubes of 10 mL per volunteer (BD Vacutainer Clot Activator tubes #367896). Blood was allowed to clot for at least 1h at RT. Samples were then centrifuged in a refrigerated centrifuge with a horizontal rotor at 1,300 x *g* for 10 min at 22ºC. Serum was collected and stored at –80°C until use. Hemolyzed samples were rejected. For the isolation of PBMCs, blood was collected in 3 heparinized tubes (BD Vacutainer #367874, 10 mL) and stored at RT until processing. Samples were diluted with PBS (1:1) and centrifuged for 10 min at 1,200 x g (RT) in SepMate™ tubes (StemCell Technologies) with density-gradient medium (Lymphoprep). The plasma was then and PBMCs were isolated by pouring them into a clean tube. Isolated PBMCs were washed twice with sterile PBS, counted, and cryopreserved in FBS (Industrial Biologicals) and 10% DMSO (Chem Cruz). All PBMC samples were stored in liquid nitrogen until use.

The neutralizing capacities of the antibodies in the samples of the volunteers were evaluated through the SARS-CoV-2 sVNT kit from Genscript (cat. number L00847-A). Assays were performed according to the instructions of the manufacturer. Briefly, serial dilutions of the serum were prepared and then incubated with the HRP-RBD reagent supplied in the kit for 30 min at 37ºC to allow the binding of neutralizing antibodies to S1-RBD. Sera and controls previously incubated with the HRP-RBD were added to the ELISA plate pre-coated with the human angiotensin-converting enzyme 2 (hACE2) protein and incubated for 15 min at 37ºC. After the incubation, samples were discarded, and plates were washed. Finally, a developing solution provided in the kit was added for 15 min at RT and then quenched with a stop solution also supplied. Plates were read at 450 nm in a microplate reader (Biotek, Ref. 1506021). The inhibition rate of HRP-RBD binding was calculated as follows: 100 * [OD450nm value of negative control - OD450nm value of sample] / [OD450nm of negative control]. Samples with a percentage of inhibition ≤30 at the lowest dilution (1:4) were assigned as seronegative with a titer of 2. A sample was considered seropositive when its titer is higher than the pre-immune titer. The methods used in this study for the detection of total and neutralizing antibodies were validated with the First WHO International Standard

Anti-SARS-CoV-2 Immunoglobulin (Cat# 20/136 and #20/268), kindly provided by PATH.

To assess the capacity of the antibodies against SARS-CoV-2 circulating variant of concern to inhibit RBD and hACE2 interaction, we performed *in-house* SARS-CoV-2 sVNT based on previous reports [1], RBD unconjugated proteins from *S1-D614G* SARS-CoV-2 (GenScript #Z03507) and Gama B.1.617.2 (GenScript #Z03613), and Omicron variant (GenScript # Z03728-1) were conjugated to HRP using the HRP Conjugation Kit - Lightning Link (#ab102890) in a 2:1 mass ratio (HRP to RBD) following the instruction of the manufacturer. ELISA 96- well plates (SPL) were pre-coated with 100ng per well of the recombinant hACE2 protein (GenScript #Z03484) in 50mL of 100 mM carbonate–bicarbonate coating buffer (pH 9.6) ON at 4°C. Plates were then washed three times with PBS - 0.05% Tween 20 and blocked with PBS - 10% FBS for 2h at RT. The HRP-RBD conjugates obtained previously were then incubated with the serum sample in a final volume of 120 µL for 1 h at 37°C. Concentration of conjugates used were as follows: 3 ng of WT SARS-CoV-2, 3 ng of B.1.1.529 and 3 ng of B.1.617.2. Then, these mixtures were added into the 96-well plates coated with hACE2 and were incubated for 1h at Room Temperature (RT). Unbound HRP-RBD were removed washing five times with PBS - 0.05% Tween 20. Then, 50 µL of 3,3',5,5'-tetramethylbenzidine (TMB – BD #555214) was added. An equal volume of 2 N $H_2SO^4$ was added to stop the reaction, and optical densities (OD) values at 450 nm were read. The antibody titer was determined as the last fold-dilution with a cut-off value over 20% of inhibition. The percentage of inhibition was defined as: [OD450nm value of negative control-OD450nm value of sample]/[OD450nm value of negative control*100]. Negative controls (corresponding to sera sample obtained before immunization) were included.

Pseudotyped virus neutralization test (pVNT) was also performed in order to evaluate the neutralizing antibodies against D614G, Gamma and Omicron variant. Antibodies with neutralizing capacities were measured using an HIV-1 backbone expressing firefly luciferase as a reporter gene and pseudotyped with the SARS-CoV-2 spike glycoprotein (HIV-1-SΔ19) as previously described[2]. Briefly, serum samples were initially diluted 1:4 in DMEM, serially diluted 1:3 up to 1:8,748 and then mixed with approximately 4.5 ng of p24 equivalents of HIV-1-SΔ19 in white 96-well plate. Plates were incubated during 1 hour at 37°C and then 100 mL of DMEM containing $1x10^4$ HEK-ACE2 cells was added to each well. Firefly luciferase activity was measured 48h later, using the Luciferase Assay System (Promega) in a Glomax®-96 microplate luminometer (Promega). Estimation of the ID50 was obtained using a 4-parameter nonlinear regression curve fit measured as the percent of neutralization determined by the difference in average relative light units (RLU) between test samples and pseudotyped virus controls as previously described. Data analyses and statistical analyses were carried out using GraphPad Prism v8.

To assess the cellular immune response, ELISPOT and flow cytometry assays were performed using PBMCs from volunteers at different times: the first dose (pre-immune); two weeks after the second dose; four weeks after the second dose; twenty weeks (or five months) after the second dose; four weeks after the booster (third) dose (**Figure 1B**). Upon thawing, cells were resuspended in fresh media in a 1:10 dilution to remove DMSO remnants from the freezing media. Then, cells were centrifuged, resuspended in fresh media, and counted in an automated cell counter (Logos Biosystems #L40001). Cells were adjusted to $6x10^6$ cells/mL and kept at 37ºC, 5% $CO_2$ for 15 min until use in the corresponding assay. ELISPOT plates containing a PVDF membrane were activated with 15 μL of 70% ethanol (Merck), washed three times with sterile 1x PBS, and then coated with human IFN-γ and IL-4 capture antibodies (1:250 and 1:125, respectively, CTL). After 2h of activation at RT, plates were washed two times with PBS and two times with PBS-Tween 20 0.05%. The stimulus included in these assays considers the use of Mega Pools (MPs) of peptides derived from SARS-CoV-2 proteins, previously described [3]. Two MPs composed of peptides from the S protein (MP-S) and the remaining proteins of the viral particle (MP-R) were used, as previously described [3]. These peptides were determined *in silico* to optimally stimulate $CD4^+$ T cells. Also, two MPs composed of peptides from the proteome of SARS-CoV-2 (CD8-A and CD8-B) were used, as previously described[3]. These peptides were determined *in silico* to optimally stimulate $CD8^+$ T cells. As positive controls, an independent stimulation was performed with 5 mg/mL of Concanavalin A (ConA) (Sigma Life Science #C5275-5MG) and with an MP of peptides derived from cytomegalovirus proteins (MP-CMV) for the stimulation of both $CD4^+$ and $CD8^+$ T cells [3]. As a vehicle control, DMSO 1% (Merck #317275) was included. A total of $3x10^5$ cells in 50 μL of media were added to each well containing 50 μL of media with the corresponding stimulus. The final concentration of each stimulus per well was 1 μg/mL (except for ConA and DMSO). Positive controls for ELISPOT assays considered $5x10^4$ cells/well instead of $3x10^5$ cells/well. For ELISPOT assays, cells were incubated for 48h at 37ºC, 5%

$CO_2$. After incubation, plates were washed 1 time with PBS and 3 times with PBS-Tween20. Then, anti-human IFN-γ (FITC) and anti-human IL-2 (Biotin) antibodies (1:1,000 and 1:1,000, respectively) were added and plates were incubated for 2h, RT. Plates were washed 3 more times with PBS-Tween 20 and then FITC-HRP and Streptavidin-AP (1:1,000) were added and plates were incubated for 1h, RT. After incubation, plates were washed 3 more times with PBS-Tween20. Then, plates were treated with the blue (15 min) and red (15 min) developer solution individually following the recommendations of the manufacturer. Plates were washed with tap water after each developer solution and allowed to dry for 24h prior to reading. To evaluate the number of T cells secreting IFN-γ, IL-4 or both, ELISPOT assays were performed with ImmunoSpot® technology (ImmnunoSpot® #hIFNgIL4-1M-10). Spot Forming Cells (SFCs) were counted on an ImmunoSpot® S6 Micro Analyzer.

To characterize the expression of activation-induced markers (AIM) by T cells, flow cytometry assays were performed. $5x10^5$ cells per well were stimulated as described for the ELISPOT assays and after 24 h of incubation with the stimulus, samples were stained. Staining was performed by incubation for 45 min at 4ºC using the reagents listed in Supplementary **Table 1**. Cells were washed twice with 200 µL of PEB buffer, fixed, and then handed to the Flow Cytometry core facility, for their acquisition in an LSRFortessa X-20 flow cytometer.

Cytokine secreted by PBMCs stimulated with MP peptides was performed on a Luminex 200, using a Milliplex MAP Mouse magnetic bead kit (Merck Millipore), following manufacturer instructions. Supernatants of samples stored at -80ºC were thawed at room temperature and diluted 1:2 before analysis. After 2 h incubation with spectrally encoded beads, coated with analyte-specific biotinylated primary antibodies, samples were incubated with streptavidin R-phycoerythrin.

**Supplementary table 1: Reagents used in Flow cytometry**

| Marker | Clone | Fluorophore | Vendor | Dilution |
|---|---|---|---|---|
| CD3 | OKT3 | Alexa Fluor 700 | BioLegend | 1:50 |
| CD4 | RPA-T4 | BV605 | BioLegend | 1:50 |
| CD8 | RPA-T8 | BV650 | BioLegend | 1:50 |
| CD14 | M5E2 | V500 | BD | 1:100 |
| CD19 | HIB19 | V500 | BD | 1:100 |
| CD69 | FN50 | PE | BD | 1:10 |
| CD137 | 4-1BB | APC | BioLegend | 1:50 |
| OX40 | BER-ACT35 | PE-Cy7 | BioLegend | 1:50 |
| Fixable Viability Dye | - | BV510 | BD | 1:1000 |

### Supplementary Figure

**Supplementary Figure 1**: **Titers of antibodies inhibiting the interaction between the S1-RBD and hACE2 in volunteers that received the booster dose**. Inhibiting antibodies were detected in serum of volunteers immunized with CoronaVac® using a surrogate Viral Neutralization Test (sVNT), which quantifies the interaction between S1-RBD and hACE2 on ELISA plates. Results were obtained from a total of seventy-seven volunteers (**A**), 36 of them were adults between 18-59 years old (**B**), and 41 of them were adults ≥ 60 years old (**C**). Data is represented as the logarithm of reciprocal antibody titer regarding the time after the first dose. Numbers above the bars show the Geometric Mean Titer (GMT), and the error bars indicate the 95% CI. A repeated measures One-Way ANOVA test assessed statistical differences to compare all times against 3$^{rd}$ dose + four weeks. **p<0.005, ****p<0.0001.

**Supplementary Figure 2**: **Changes in activation-induced markers (AIMs) expression in CD8$^+$ T cells through flow cytometry and number of IFN-g-secreting cells upon stimulation with Mega Pools of peptides derived from SARS-CoV-2 measured after the booster dose of CoronaVac®.** AIM$^+$ CD8$^+$ T cells were quantified in peripheral blood mononuclear cells of volunteers that received a booster dose of CoronaVac® twenty weeks after the second dose by flow cytometry, upon stimulation with mega-pools of peptides derived from SARS-CoV-2 proteins. The percentage of activated AIM$^+$ CD8$^+$ T cells CD69$^+$, CD137$^+$ were determined upon stimulation for 24h with MP-S+R in samples obtained at pre-immune, two weeks after the second dose, four weeks after the second dose, twenty weeks the second dose, and four weeks after the booster dose. Data from flow cytometry was normalized against DMSO and analyzed separately by a Friedman test against the booster dose. Results were obtained from a total of forty volunteers (**A**), twenty-one were of them were adults between 18-59 years old (**B**), and nineteen of them were ≥ 60 years old (**C**). Changes in the secretion of IFN-γ were quantified as the number of Spot Forming Cells (SFCs) in peripheral blood mononuclear cells of volunteers that received a booster dose of CoronaVac® 20 weeks after the second dose. **D-F**. Data was obtained upon stimulation with MP-S+R for 48h in samples obtained at pre-immune, two weeks after the second dose, four weeks after the second dose, twenty weeks the second dose, and four weeks after the booster dose. Results were obtained from a total of 40 volunteers (**D**), 21 were of them were adults between 18-59 years old (**E**), and 19 of them were ≥ 60 years old (**F**). Data from ELISPOT were analyzed separately by Friedman test against the booster dose with not statistically differences.

**Supplementary Figure 3: IL-2 and IFN-γ secretion is induced in PBMC stimulated with Mega-Pools of variant of SARS-CoV-2 four weeks after the booster dose with CoronaVac®.** Changes in cytokine secretion were determined by Luminex®. Data was obtained upon stimulation of PBMCs with MP against protein S of WT, Delta and Omicron variant of SARS-CoV-2 for 20h in samples obtained at four weeks after the booster dose. Results were obtained from a total of twenty volunteers (**A**) IL-2 secretion and (**B**) IFN-γ Data shown represent means + 95%CI. A repeated measures One-Way ANOVA test assessed statistical differences of the GMT to compare each variant against D614G.

**Supplementary Figure 4: Quantification of circulating neutralizing antibodies live SARS-CoV-2 variants in volunteers that received the booster dose of CoronaVac®**. **A.** Neutralizing antibodies were detected in serum of nineteen volunteers that received a booster dose of CoronaVac® twenty weeks after the second dose, using a conventional Viral Neutralization Test (cVNT), which quantifies the reduction of cytopathic effect (CPE) in Vero E6 cells infected with SARS-CoV-2 variants D614G and Delta (Chilean isolates). **B.** Quantification of the titers of neutralizing antibodies from nineteen volunteers vaccinated with CoronaVac®, four weeks after the booster dose. Data are expressed as the reciprocal of the highest serum dilution preventing 100% cytopathic effect, the numbers above each set of individual data points show the Geometric Mean Titer (GMT) and the error bars indicate the 95% CI. **D.** Seropositivity rate of neutralizing antibodies are shown for each time point analyzed. Numbers above the bars show the percentage of seropositivity rate in the respective graphs. A Wilcoxon-T test was performed to assessed statistical differences of the GMT to compare between groups. *p<0.05; ***p < 0.001.

**Supplementary Table 2: Seropositivity rates, Geometric Mean Titer (GMT) of circulating neutralizing antibodies against SARS-CoV-2 RBD of D614G and Delta variant.**

| | Variant | D614G | Delta (B1.617.2) |
|---|---|---|---|
| | Indicators | 3rd dose + 4 weeks | 3rd dose + 4 weeks |
| cVNT | Seropositivity n/N | 19/19 | 16/19 |
| | (%) | 100 | 84 |
| | GMT | 128.0 | 14.3 |
| | 95% CI | 60.6-270.2 | 8.2-25.1 |

*GMT: Geometric mean titer.*

**Supplementary Appendix**

**1. CoronaVac03CL Study Group / Sept 14, 2021**

**Center CL1: Áreas Ambulatorias Marcoleta - Pontificia Universidad Católica de Chile.**

1. Álvaro Miguel Rojas Gonzalez,
2. María Soledad Navarrete Bello,
3. Constanza Belén Del Rio Solis,
4. Dinely Valeska Del Pino Lavín,
5. Natalia Elizabeth Aguirre Concha,
6. Franco Vega Farías,
7. Acsa Raquel Salgado Ovalle,
8. Thomas Quinteros,
9. Alma Muñoz,
10. Patricio Astudillo,
11. Monique Nicole Le Corre.

**Center CL2: Clínica San Carlos de Apoquindo - Red de Salud UC-Christus**

1. Marcela Potin Santander
2. Sofía Aljaro Ehrenberg
3. Sófia López Coloma
4. Tania Weil Valjalo
5. Gema Pérez Alarcón
6. Melan Peralta Kong
7. Consuelo Zamanillo Moreira

**Center CL4: Clínica Los Andes - Universidad de Los Andes**

1. Paula Guzmán Merino,
2. Francisca Aguirre Boza,

3. Aarón Cortés Rojas,
4. Luis Federico Bátiz,
5. Javiera Francisca Pérez Velásquez,
6. Karen Pamela Apablaza García,
7. Lorena Yates Barsotti,
8. María de los Ángeles Valdés Valdés,
9. Bernardita Hurtado,
10. Veronique Venteneul,
11. Constanza Astorga.
12. Maria Francisca Bossans
13. Ximena Correa
14. Pilar Navarro
15. Javiera Lagas

**<u>Center CL5</u>: Clínica Alemana - Universidad del Desarrollo**

1. Paula Andrea Muñoz-Venturelli,
2. Pablo Agustín Vial,
3. Andrea Ingrid Schilling Redlich,
4. Daniela Pavez Azurmendi,
5. Inia Andrea Pérez Villa,
6. Amy Lisa Riviotta,
7. Francisca Gonzalez Mc Cowley,
8. Francisca Pilar Urrutia Goldsack,
9. Alejandra Isabel Del Río Weldt,
10. Claudia Andrea del Carmen Asenjo Lobos,
11. Bárbara Paulina Vargas Latorre,
12. Francisca Valentina Castro Fuentes,
13. Alejandra Patricia Acuña Rogel,
14. Javiera Constanza Gúzman Cancino,
15. Camila Alejandra Astudillo Griffiths.
16. Camila Portilla Fuentes
17. Paulina Bustos Alarcón,
18. Carlos Delfino Garay

**<u>Center CL6</u>: Hospital Clínico Félix Bulnes - Universidad San Sebastián**

1. Carlos M Pérez,
2. Pilar Espinoza,
3. Andrea Martínez,
4. Marcela Arancibia,
5. Harold Romero,
6. Cecilia Bustamante,
7. María Loreto Pérez,
8. Natalia Uribe,
9. Viviana Silva,
10. Bernardita Morice,

11. Marco Pérez,
12. Clara Alvarado.

**Center CL7: Hospital Dr. Gustavo Fricke - Universidad de Valparaíso**

1. Marcela González,
2. Nataly Martínez,
3. Camila Molina,
4. Juliette Sánchez.

**Center CL8: Hospital Carlos Van Buren- Universidad de Valparaíso**

1. Daniela Fuentes Hulse,
2. Yolanda Calvo Toro,
3. Mariela Cepeda Corrales,
4. Rosario Lemus Manzur,
5. Constance Marucich Baeza,
6. Cecilia Cornejo Beas

**Center CL9: Complejo Asistencial Dr. Sótero del Rio**

1. Paulina Donato Inostroza,
2. Martin Lasso Barreto,
3. María Iturrieta Meléndez,
4. María Acuña Schlegel,
5. Ada Cascone Scarpati,
6. Raymundo Rojas Araya,
7. Camila Sepúlveda Contreras,
8. Mario Alex Contreras,
9. Yessica Campisto Sanhueza,
10. Pablo González Sanhueza,
11. Zoila Quizhpi Mejias,
12. Mariella Lopez García,
13. Vania Pizzeghello Salfate,
14. Stephannie Silva Monsalve.

### 2. Members of the Independent Data Safety Monitoring Committee.

Luis Delpiano, MD, Pediatric Infectologist, Hospital San Borja Arriarán, Santiago, Chile.

Macarena Lagos, MD, Immunologist, Clínica Las Condes and Hospital Padre Hurtado, Santiago, Chile.

Gloria Icaza, MD, Epidemiologist and Statitician, Universidad de Talca, Talca, Chile.

Leonardo Chanqueo, MD, Adult Infectologist, Hospital San Juan de Dios, Santiago, Chile.

Mónica Imarai, PhD, Universidad de Santiago, Santiago, Chile.
