## Supplementary figures and images for "A booster dose of an inactivated SARS-CoV-2 vaccine increases neutralizing antibodies and T cells that recognize Delta and Omicron variants of concern"

### Supplementary Figures

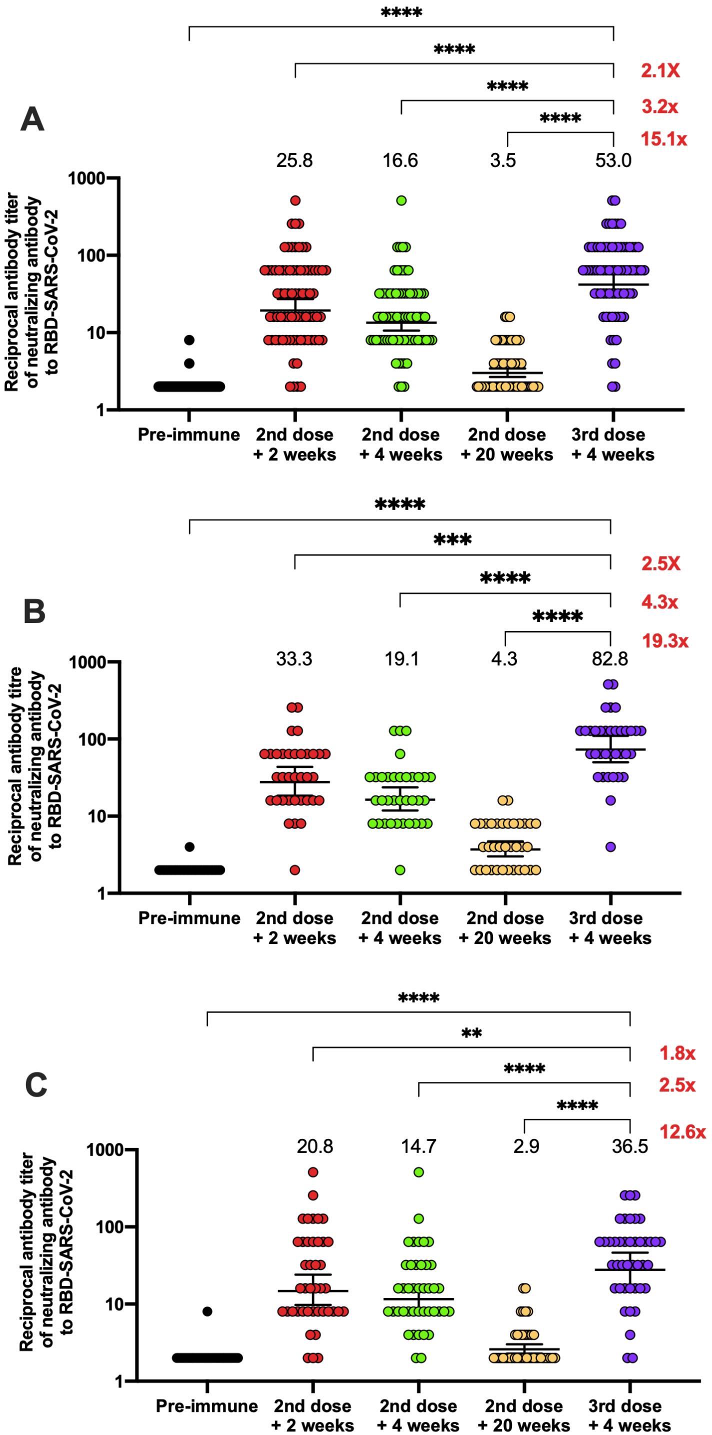

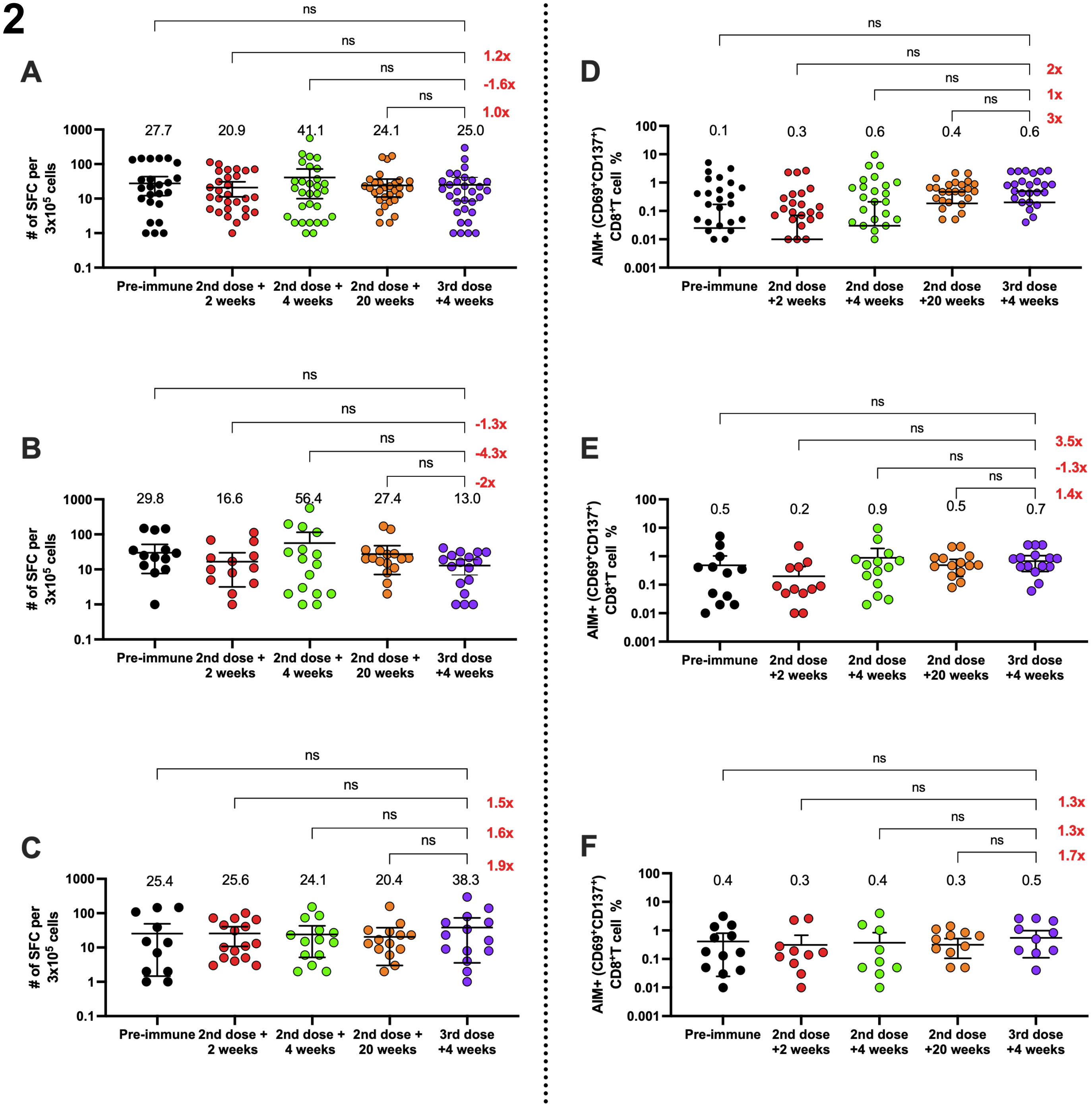

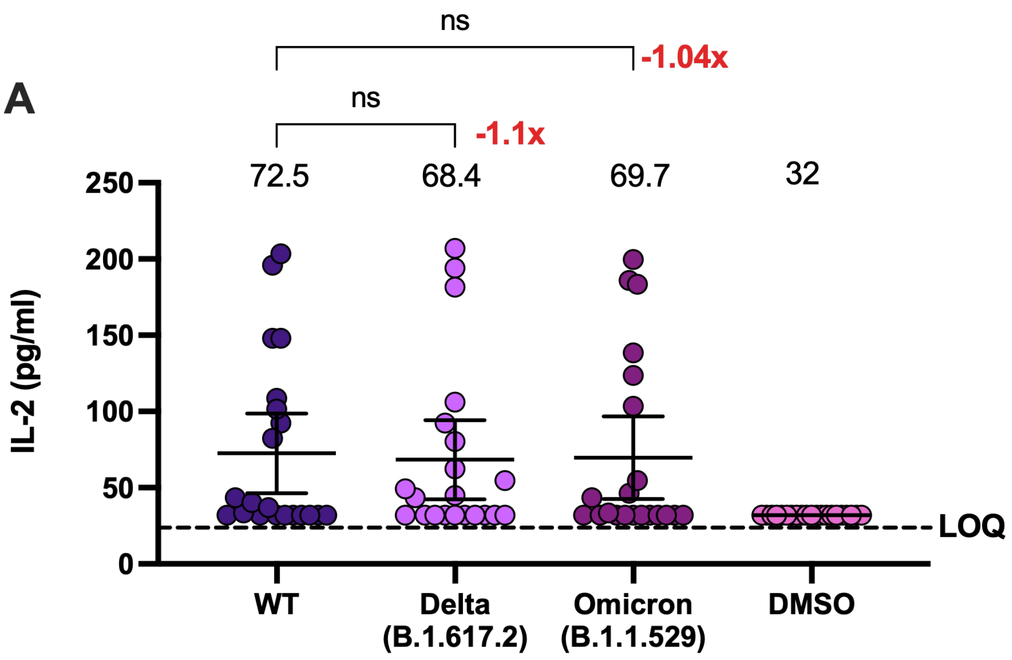

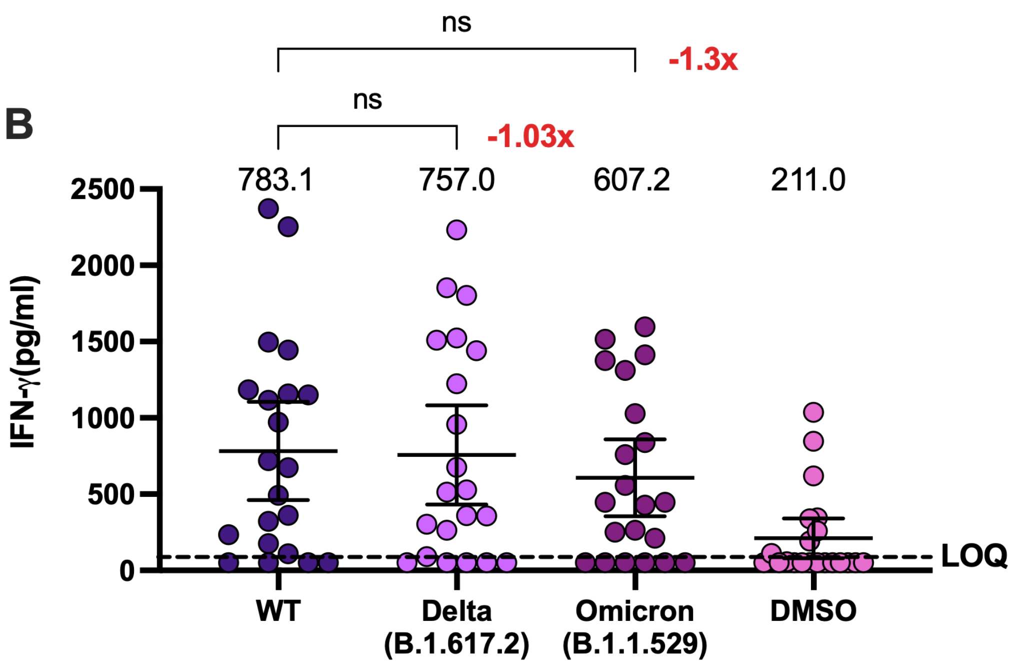

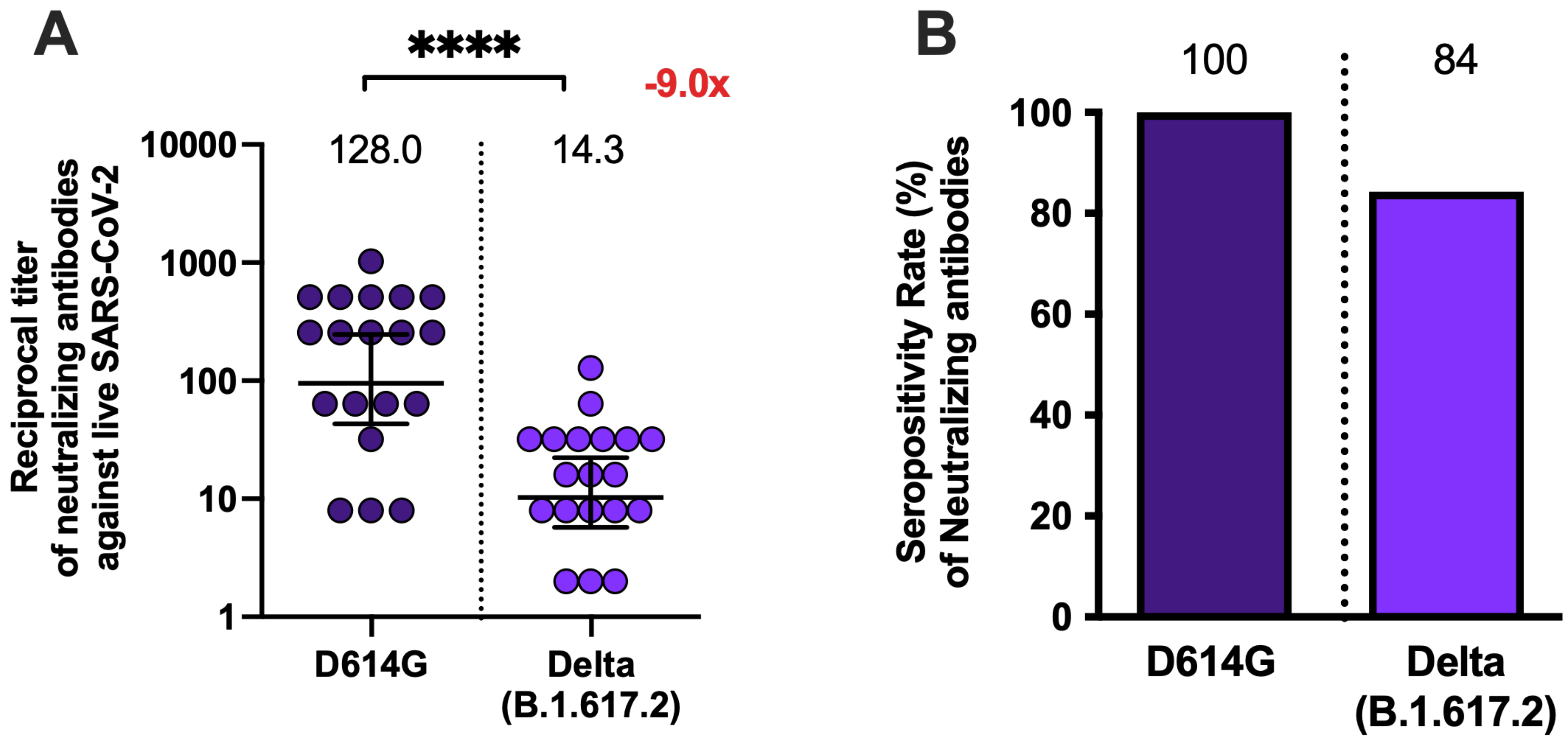
